# A systematic review of utilisation of diurnal timing information in clinical trial design for Long QT syndrome

**DOI:** 10.1101/2020.07.23.20160978

**Authors:** Lydia M Seed, Timothy J Hearn

## Abstract

Diurnal oscillations in human cardiac electrophysiology are thought to be under the control of the endogenous circadian clock. The incidence of arrhythmic events in patients with Long QT syndrome (LQTS) varies diurnally. The diurnal variation in QT interval has previously been identified as a potential for error in clinical trials which utilise ECG measurement. We performed a systematic review of clinical trials for LQTS to identify practice around specification of timing information for point electrocardiogram (ECG) measurements, analysis of continual ECG recordings ≥24 hours, and drug delivery. Despite guidelines having been issued around the analysis of 24-hour ECG recordings, we identify a lack of usage of detailed time of day information in trial design for LQTS studies, which has the potential to affect the interpretation of the results of drug trials. We identify that, in contrast, clinical trials for QT prolonging drugs demonstrate increased incorporation of time of day information of both QT analysis and drug dosing. We provide a visual portal to allow trial designers and clinicians to better understand timing of common cardiac-targeting drugs, and to bear this concept in mind in the design of future clinical trials.

## Introduction

Human physiology is synchronised to the environmental day-night cycle by an endogenous circadian clock. This clock is present as a transcriptional oscillatory network within individual tissues. Circadian rhythms in cardiovascular physiology have been the subject of much attention recently (1), featuring on various popular media outlets (2).

Circadian cardiac electrophysiology is driven by both the central circadian pacemaker, located in the suprachiasmic nucleus (SCN), and the local cardiac clock, which drives a distinct pattern of diurnal and circadian transcription in the heart (3). The SCN clock drives the circadian regulation of the autonomic nervous system, thus generating circadian variations in cardiac autonomic tone, which alters ionic conductance (4). The local cardiac clock drives circadian ion channel remodelling (5,6). The SCN clock may entrain the local cardiac clock, driving circadian oscillations in ion channel expression in the heart, but further research is needed to clarify the synchronisation of the two clocks (3) and resulting tissue specific rhythm.

Diurnal variation in cardiovascular physiology, such as nocturnal bradycardia and decrease in blood pressure (7), has long been established in the literature. Cardiac electrophysiology exhibits diurnal variation that manifests in altered electrocardiogram (ECG) waveforms: during the night, the duration of the corrected and uncorrected QT interval increases, reflecting slower ventricular repolarisation (8,9) (Figure 1 A).

**Figure 1.**
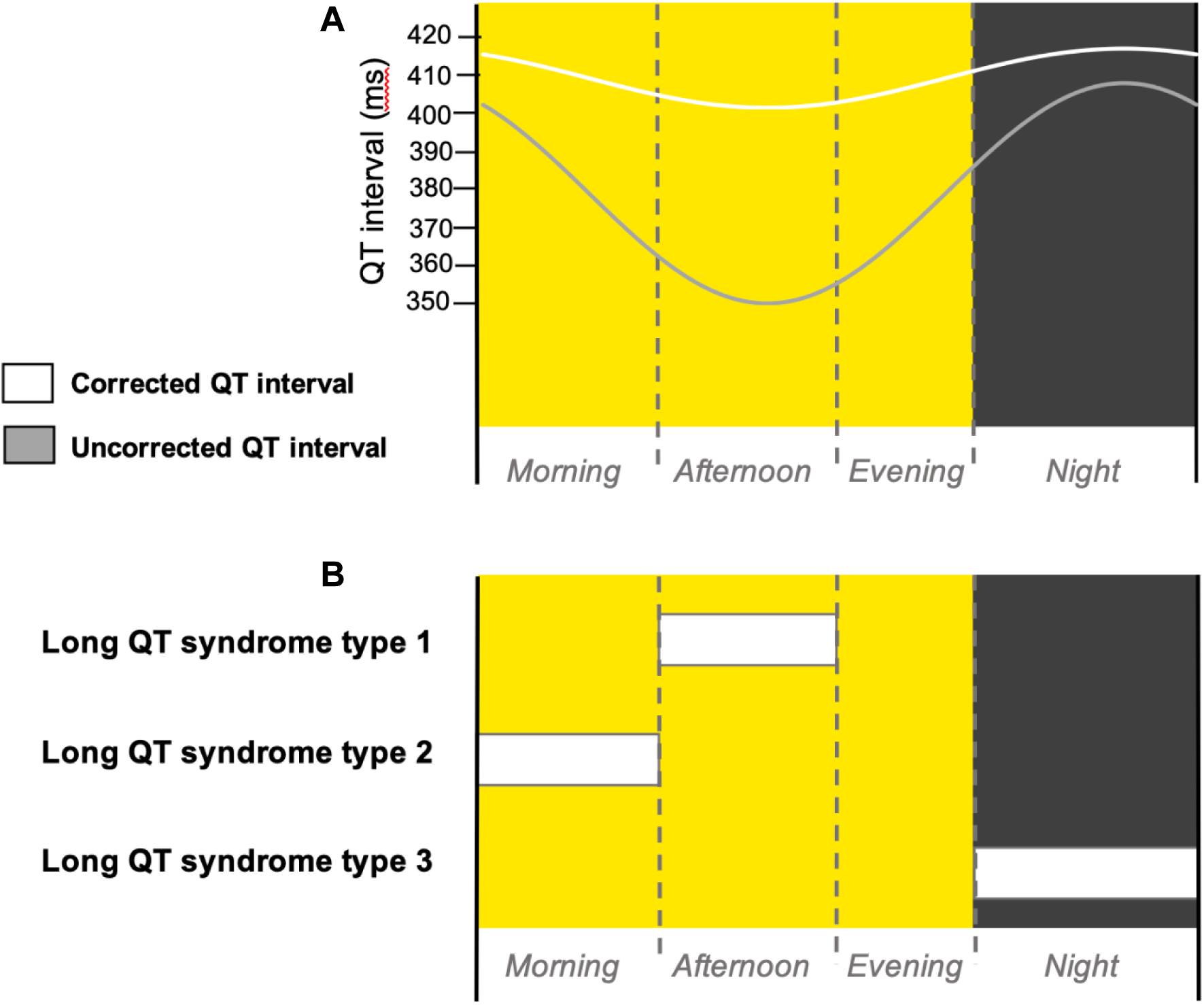
Diurnal variation in QT interval and cardiac events in LQTS patients. **(A)** Schematic cartoon diagram of the diurnal variation of corrected (white) and uncorrected (grey) QT intervals (9,15) over a long day photoperiod diurnal cycle. **(B)** Schematic summary of the times of day when patients with different subtypes of LQTS most frequently experience cardiac events (11–14). Yellow background indicates daytime and grey background indicates night-time. Dotted line indicates the transition between morning, afternoon and evening phases of the diurnal cycle.

Long QT syndrome (LQTS) is an inherited arrhythmia characterised by a prolonged QT interval – the time from the beginning of the QRS complex to the end of the T wave on an ECG. This electrical instability of prolonged ventricular repolarisation, in an otherwise structurally normal heart, can trigger life-threatening heart rhythm abnormalities, such as ventricular fibrillation, polymorphic ventricular tachycardia and Torsades de Pointes, which can lead to cardiac arrest. Consequently, LQTS is associated with an increased risk of sudden cardiac death (10). The incidence of cardiac events in LQTS patients displays a circadian basis, specific to each LQTS subtype (Figure 1 B). It is recognised that the incidence of cardiac events in patients with LQTS type 1 is highest during the afternoon (11), LQTS type 2 (LQT2) is highest during the morning (11) and LQTS type 3 (LQT3) is highest during the night (12–14).

Therefore, LQTS is an excellent candidate for diseases where the circadian clock contributes to the pathology, as both the underlying electrocardiographic phenotypes and the risk of arrhythmic events demonstrate diurnal oscillations.

Drug-induced QT prolongation, caused by a wide range of commonly prescribed drugs, can trigger similar life-threatening arrhythmias, namely Torsades de Pointes, in patients who are otherwise not predisposed. The QT-prolonging effects of drugs are a common evaluation feature during clinical trials and are often included as secondary outcomes for evaluating drug safety.

The diurnal oscillations in cardiac electrophysiology need to be taken into account when reviewing the effectiveness of clinical interventions, for example in clinical trials that use such oscillating metrics as outcomes of intervention success. However, current clinical trial designs are inconsistent in respect of the time of day of measurement of cardiac electrophysiological parameters using ECG, or for time of day of drug delivery. Here we analyse the current clinical trial literature to identify the extent to which trials carried out in LQTS patients, or that evaluate the QT-prolonging effects of drugs administered, are currently recording or incorporating diurnal information into their trial strategies, and we suggest guidelines based upon these findings.

## Materials and methods

### Search strategy and selection criteria

To assess the inclusion of diurnal reporting in ECG measurements in clinical trials for LQTS, or that evaluate QT-prolonging effects of drugs, a local database of clinical trails was built (Supplemental Table 1). Trial information was extracted from the European and United States clinical trials databases by searching “Long QT syndrome” and “Long QT” in https://www.clinicaltrialsregister.eu/ctr-search/search and “Long QT syndrome” in the ‘Condition or disease’ search bar and “Long QT” in the ‘Other terms’ search bar in https://clinicaltrials.gov/ct2/home, with an exact cut-off date of 21^st^ June 2020. This was expanded by the search algorithm to include prolonged QT interval. Trials were first deduplicated and then excluded based on the following criteria: not providing any detailed information on how the clinical trial was conducted; not recording any form of ECG; or not providing detail on the method of ECG recording or analysis (Figure 2). The remaining trials were interrogated to determine which method(s) of ECG recording were used: point – an ECG recording for <1 hour, usually using a standard 12-lead machine – or continual – an ECG recording for ≥24 hours, usually using a Holter monitor. It was then determined whether time of day information was reported in the recording of point ECGs and in the analysis of continual ECGs. We defined the time of day reporting for point ECG recordings as noting the time of ECG recording (either specifically or in more vague terms such as “in the morning”), or taking multiple ECG recordings evenly spaced over 24 hours, or recording and analysing ECGs in a time-matched manner. We defined time of day analysis of continual ECG recordings as separately analysing daytime and night-time data. Where applicable the time of day information was also recorded for drug administration during a trial, using the same criteria for point ECGs. We decided that directions for drug administration to achieve a persistent drug dose, such as “twice a day” or “every 6 hours” was not sufficient time of day information.

**Figure 2.**
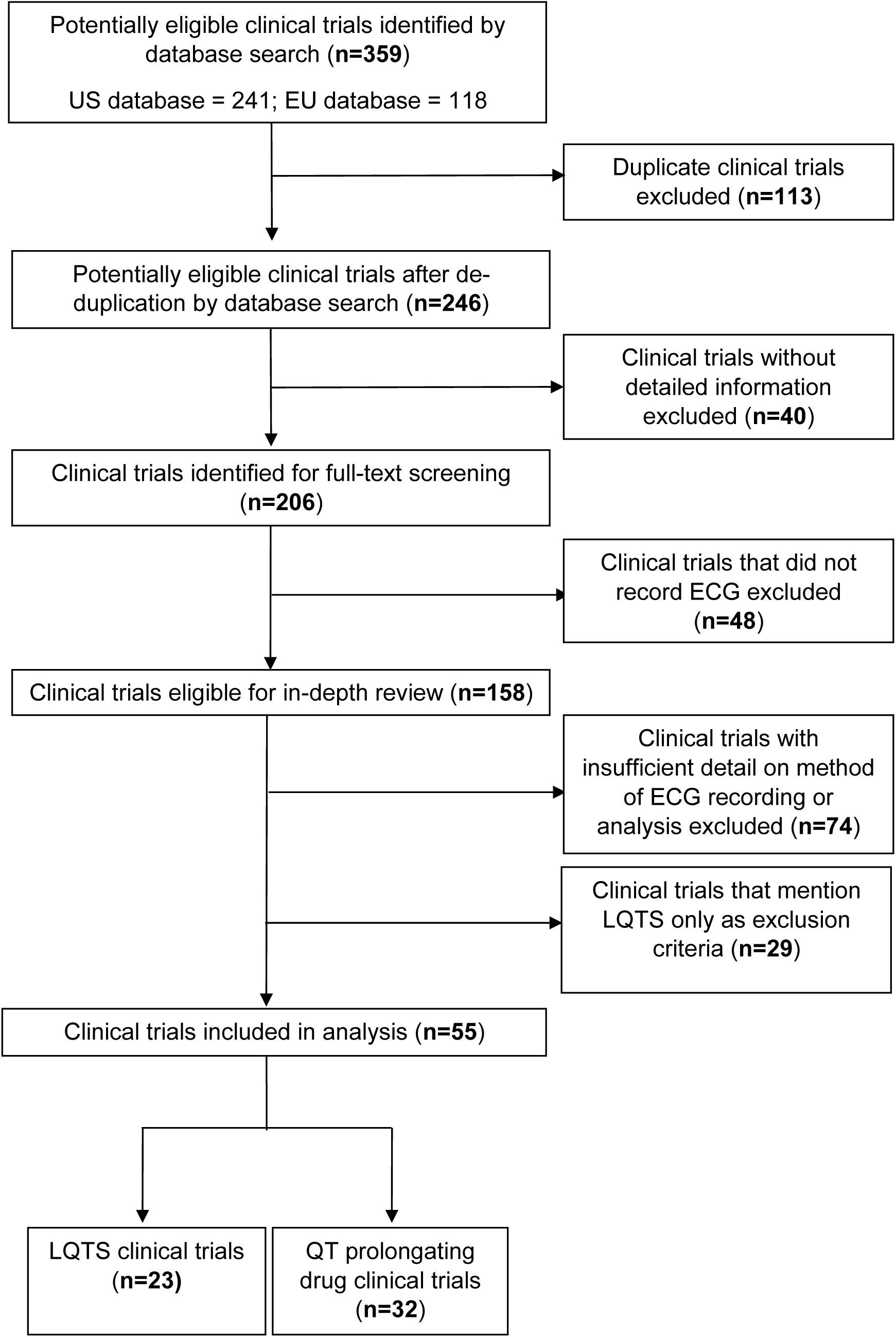
Flowchart outlining the protocol for trial selection in this study. Trials were downloaded from the EU (https://www.clinicaltrialsregister.eu) and the US (https://clinicaltrials.gov/) registers.

### Statistical analysis

All data were plotted and analysed in R. Statistical analysis was performed using the inbuilt functions for chi-squared [chisq.test()] with Yate’s correction or Fisher’s exact test [fisher.test()]. Chi-squared statistic, degrees of freedom and p-values are given in the text. Odds ratios (OR) are given for Fisher’s when p-values correspond to 2×2 contingency tables.

## Results

### There is variable inclusion of time of day information for ECG measurement or analysis in LQTS clinical trials

Following searching the US and EU clinical trials databases and removing duplicates, 246 potentially eligible clinical trials were identified. Subsequently, 162 trials were excluded due to lack of providing sufficient information, recording an ECG or detailing the method of ECG recording as demonstrated in Figure 2. A total of 55 clinical trials were included in the analysis, 23 which were clinical trials for LQTS and 32 which were trials for prolonged QT interval. The LQTS trials could be further divided into trials which were interventional, involving some form of pharmaceutical challenge, or observational in nature (Figure 3 A). We first identified whether the trials conducted point ECG recordings or performed continuous ECG recordings for ≥24 hours. The majority of LQTS trials (47%) recorded only point ECG measurements, whereas 13% recorded only continual ECGs. Both point and continual ECG recordings were performed in 14% of clinical trials, and the type of recording was not significantly different in interventional compared to observational studies (χ^2^ = 1.383, df = 2, p-value = 0.501) (Figure 3).

**Figure 3.**
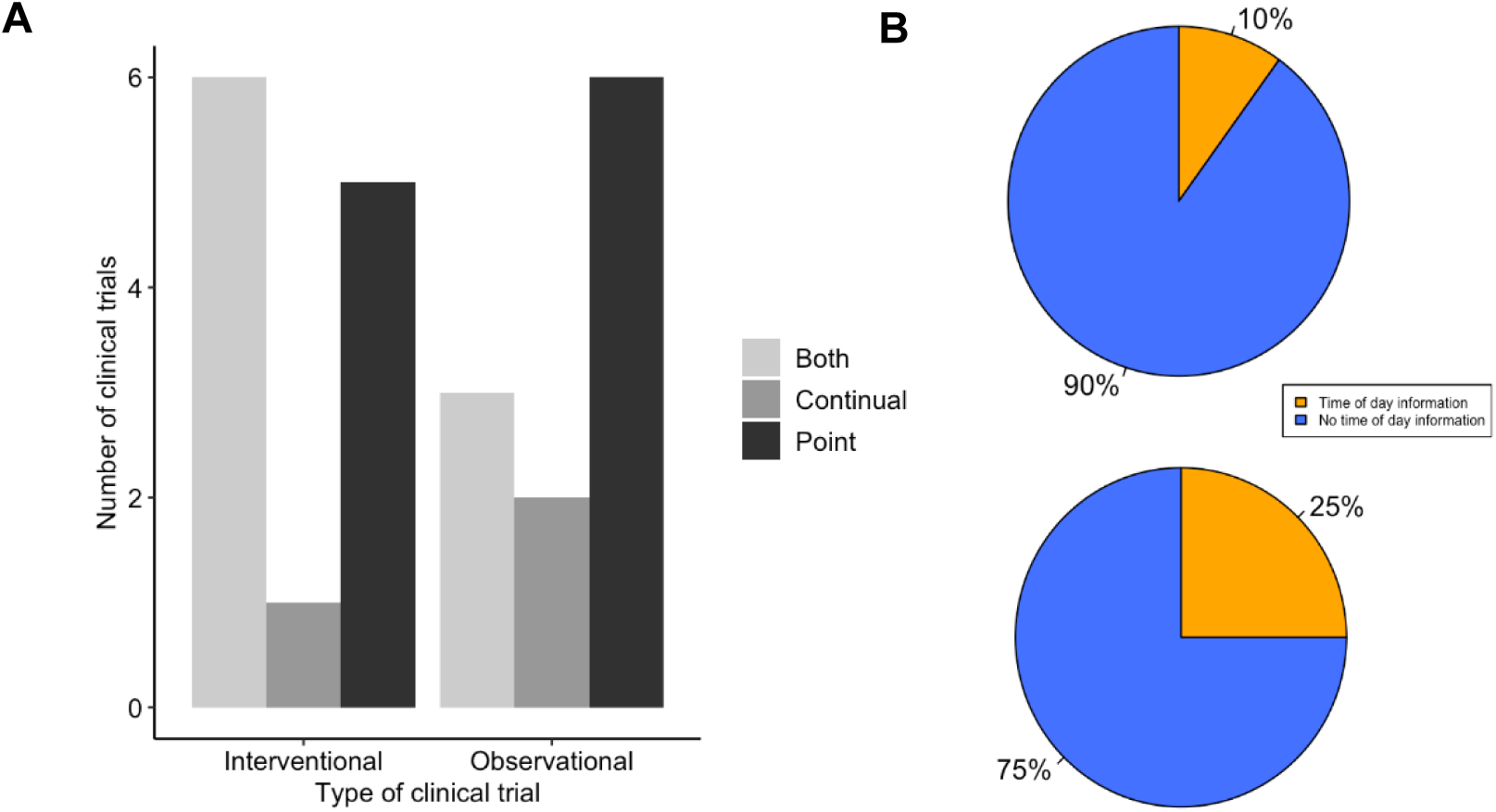
LQTS trials make low use of time of day information for ECG measurement. **(A)** Method of ECG recording used for LQTS clinical trial outcomes. Trials are grouped together as either interventional or observational studies. Trials were scored for whether outcomes reported ECG measurements determined by point ECG measurement (black), continual using Holter monitor (dark grey) or outcomes that used both point and continuous measurements (light grey). **(B)** Distribution of LQTS trials that accounted (orange) or did not account (blue) for time of day information when (top) recording or analysing point ECGs and (bottom) analysing continual ECGs.

The uncorrected QT interval can change by up to 50ms (9), and the corrected QT interval can vary by up to 10ms (15), depending upon the time of day of recording. We looked within the trial protocols for methodology on ECG recordings, to assess to what extent time of day information was recorded for the point recordings and whether it was used in the analysis of continual ECG measurements. We found that 90% (n=18) of trials which used point recordings did not have any mention of the time of day of ECG recording (Figure 3B). This contrasted to 75% (n=9) of trials that used continual ECG recordings which did not use specified time windows in their analysis and instead used average daily values (Figure 3B). This indicates that the majority of LQTS trials do not include time of day information for ECG recording or analysis. Moreover, there is not a significant difference between the use of timing information between point and continual ECG measurements (Fisher’s P value = 0.338 OR = 0.346).

It is important when taking physiological measurements that oscillate diurnally that the timing of pharmaceutical intervention is also taken into account. We observed that there was no difference in the use of point or continual ECG recordings to measure QT interval in trials that utilised drug treatment compared to those that did not (χ^2^ = 1.711, df = 2, p-value = 0.425) (Figure 4A). Many drugs are known or predicted to have different pharmacokinetics at different times of day, and therefore will influence physiology discretely based on time of dosing. We looked in the subset of LQTS trials that administered a drug to identify if the drug was dosed at a known time of day (Figure 4B). We found that only 2 trials recorded the time of day of drug administration. We analysed time of day information for ECG recording and drug administration together and found that no trials detailed time of day information for both ECG recording and drug delivery (figure 5 B).

**Figure 4.**
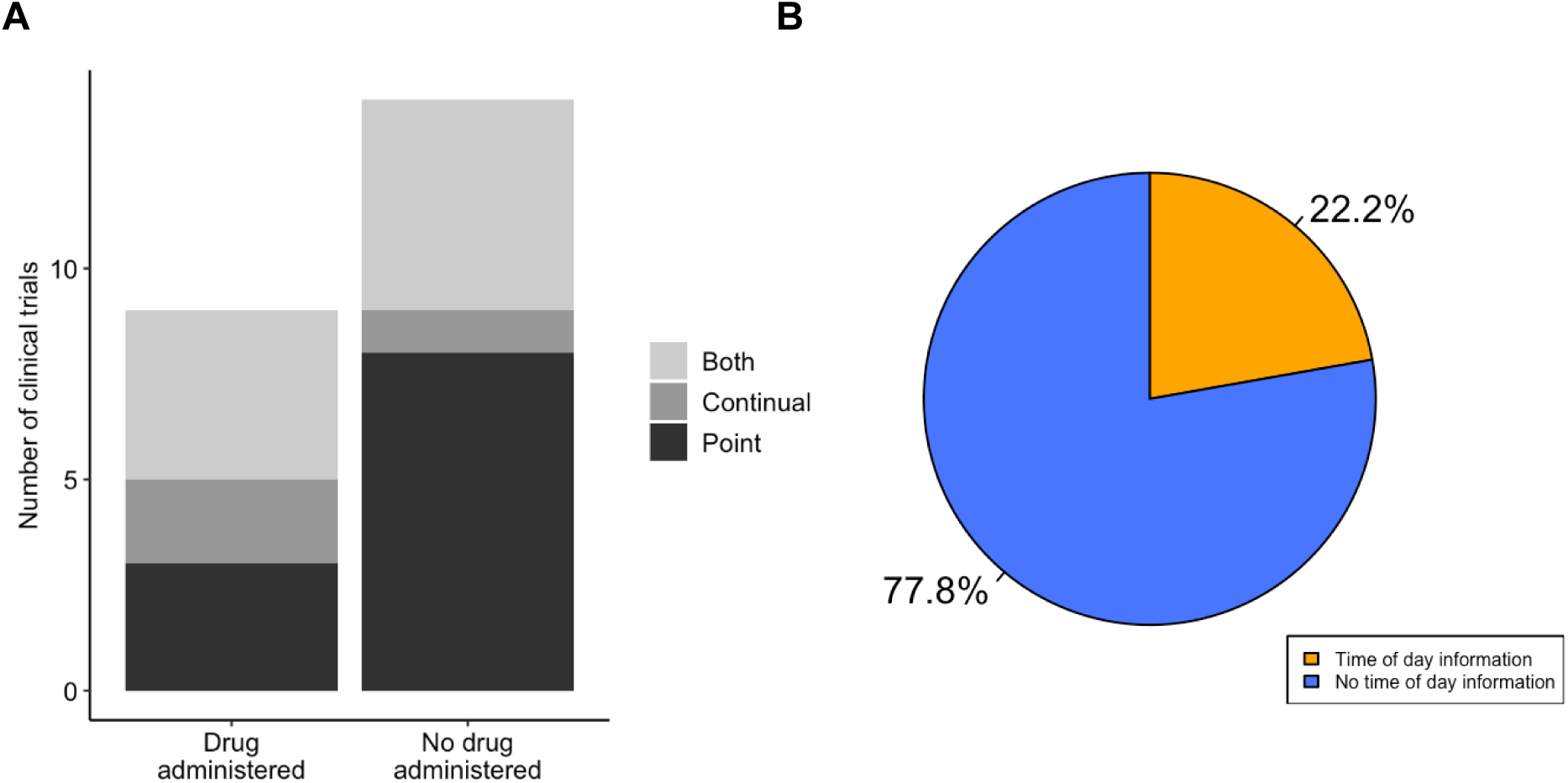
LQTS drug studies do not show enhanced integration of diurnal information. **(A)** Method of ECG recording for LQTS clinical trials that did and not administer a drug. Trials were scored for whether outcomes reported ECG measurements determined by point ECG measurement (black), continual using Holter monitor (dark grey) or outcomes that used both point and continuous measurements (light grey). **(B)** Distribution of LQTS trials that administered a drug that accounted (orange) or did not account (blue) for time of day information in drug delivery.

**Figure 5.**
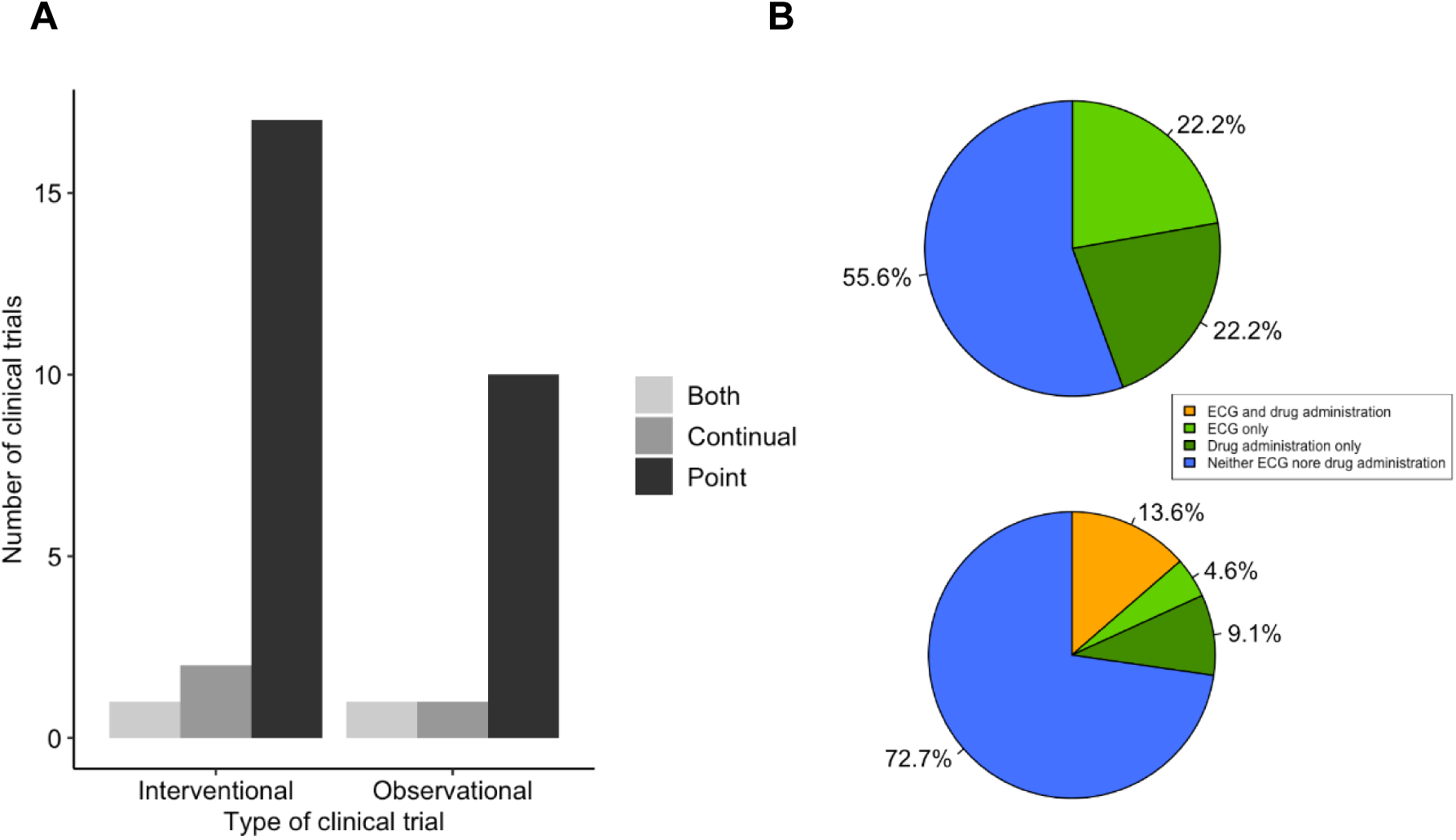
QT-prolongation drug trials integrate drug and ECG timing to a low level. **(A)** Method of ECG recording for QT prolongation drug clinical trials. Trials were scored for whether outcomes reported ECG measurements determined by point ECG measurement (black), continual using Holter monitor (dark grey) or outcomes that used both point and continuous measurements (light grey). **(B)** Integration of both drug and ECG timing in clinical trials for LQTS (top) or QT-prolonging drugs (bottom). Trials that take into account timing information are highlighted for: both ECG and drug delivery (orange), only ECG measurement / analysis (light green) and only drug administration (dark green).

### Trials for QT-prolongation effects include some integration of time of day information for ECG and drug administration

Given the difference in clinical trial design with respect to time of day information for ECG measurements and drug administration observed in the LQTS studies, we interrogated the trials for QT-prolongation effects, or “induced-long QT” effects, of drugs to identify whether similar differences in clinical trial design could be observed. The 33 trials that satisfied the criteria for inclusion in the QT-prolongation effects group contained a greater number of studies that only contained point ECG measurements, and this was significantly enriched compared to the LQTS trials group (χ^2^ = 9.986, df = 2, p-value = 0.007; Figure 5A). The use of point ECG measurements increases the importance of recording timing information, as the analysis cannot be retrospectively performed. We analysed the trials in the QT-prolongation effects group to identify whether they included timing information for ECG measurements or analysis, drug administration, or both. There was no significant difference in the number of trials that included timed ECG measurements compared to the LQTS trials (χ^2^ = 0.010, df = 1, p-value = 0.922). Moreover, there was no significant difference in the number of trials that included timed point ECG measurements (χ^2^ = 7.59×10^−32^, df = 1, p-value = 1). Within the LQTS trials that utilised drug treatment, no overlap was observed between the trials that detailed the time of day for ECG measurement and the trials that detailed the time of day for drug delivery (Figure 5B). In contrast, 13.6% (n=3) of trials in the QT-prolongation effects cohort detailed the time of day for both ECG measurement and drug delivery (Fisher’s p-value = 0.035). Further, only a single trial detailed the time of day for ECG measurements only. Overall, the QT-prolongation effects trials contained a similar proportion of trials that detailed time of day of drug delivery (22.2%) compared to the LQTS trials that administered a drug (22.7%).

### A tool to aid identification of optimal time window of measurements or analysis for clinical trial design

Previous studies have highlighted the importance of ensuring that drug timing information in chronotherapeutics is understood and used sensibly at all levels of practice (16). Over what interval is it important to take or analyse ECG measurements in clinical trials? Typically, studies are performed in the daytime for point measurements whereas 24hr Holter monitor studies have the advantage of being able to look at QTc over any time interval. However, the majority of trials do not distinguish between morning and evening measurements, in part due to current guidelines for best practice as set out in ICH E14 (17).

There are few results available in standardised format for the LQTS clinical trials. The 2 available results demonstrate the 2 extremes of practice: NCT02300558 reports primary outcomes based on the use of continuous ECG measurement to ascertain effects either in the day, night or across the entire diurnal cycle; whereas 2009-011819-20 reports point ECG measurements at an unspecified time. The reporting of only the daytime analysis as a significant outcome in NCT02300558 highlights the importance of use of this time window, compared to average QTcF measurements. However, this window may change depending on the diurnal pharmacokinetic profile of the drug being administered in the trial. Therefore, although it is optimal to take ECG measurements during the afternoon – the trough of the QT/QTc diurnal rhythm, to minimise the noise due to diurnal variation – this may not be the best window to ascertain the effects of the drug. This is especially pertinent when the time of day of cardiac events of the individual LQTS subtypes is considered. Therefore, both the diurnal variations in pharmacokinetics of the drug and the QTc estimate need to be considered, in line with ICH E14 guidance.

Based on these observations, we suggest criteria for recording point ECG measurements in drug trials that account for the diurnal variations in both cardiac electrophysiology and the pharmacokinetics of the drugs administered. Similar criteria can be applied to any physiological outcome that displays diurnal variation. These fit into CONSORT guidelines 4b, 5 and 6a (18). From the general descriptions of temporal information in the trials we have reviewed, we identify that it is difficult for researchers and trials co-coordinators to standardise this information. Therefore, we have designed a tool that can identify the most appropriate diurnal window in which to take physiological measurements, such as point ECGs, depending on the drug administered in the trial. The tool uses the predicted phase of the targets of drugs with short half-lives deposited in circaDB (19). We have incorporated the time of maximum peak to ensure that the relevant physiology is observed and reported when clinical trial analysis is conducted. This tool is available online at https://www.comparativechrono.org/tools.

## Conclusion

We assessed the extent to which diurnal reporting is included in clinical trials for LQTS that use either corrected or uncorrected QT measurement as an outcome of the trial. We noted that the majority of trials do not note the time of day of recording for point ECG recordings, but trials which utilise 24 hr Holter monitors make use of this information to a greater extent.

We noted that in LQTS trials that administer a drug there were no trials that recorded both time of ECG point recording and time of drug delivery. When we compared trials in this cohort to trials that were investigating the QT-prolongation effects of a drug as a primary outcome we found similar results. However, some trials in the QT-prolongation effects cohort noted the time of day of both QT interval measurement and drug delivery. All of the sample sizes were small due to the small number of trials that were eligible for inclusion.

Most studies use QTcF and QTcB to avoid the problem of strong differences in day/night QT interval. However, there is a circadian basis to QT change itself, and this can be seen in the corrected intervals, with a magnitude of around 40 ms. Many trials use cut-offs of 10 ms or 30 ms (e.g. 2008-003732-38) for reaching a threshold difference, which is within the magnitude of diurnal change that is typically observed. Therefore, without detailing time of day of the ECG measurement or analysis this observation could be wrongly interpreted as the effect of a clinical intervention investigated in the trial rather than the result of daily changes in cardiac electrophysiology.

As a result of ICH E14 (17), many trials now include timing information for matched measurements, some including time-matched or pre-dose baseline, but the timing is never referenced to a specific time of day. Although measurements are time-controlled between the groups of participants receiving a placebo and the drug, they appear not to be time-controlled between individual participants within each group, reducing its relevance for considering differences for the QT interval.

Diurnal analysis might not be important for many studies; however, it is crucial for the effective design of clinical drug trials when both the underlying physiology and the drug pharmacokinetics oscillate over a 24-hour period. While the growing number of QT-prolongation trials incorporating timing information for both QT interval measurements and drug administration in their protocols is promising, our finding that few LQTS drug trials incorporate diurnal information in trial design and analysis is concerning. We express this concern as QT intervals are widely used to assess patient safety in clinical drug trials. If QT intervals are measured *ad hoc*, measurements may be confounded by the effects of the diurnal variations in both cardiac electrophysiology and pharmacokinetics of the drugs administered. These factors need to be accounted for when measuring QT intervals as a clinical outcome. A consideration for future work could be to identify the inclusion of timing information into the thousands of trials that use QT measurements as clinical outcomes.

We noted that there is no standardisation in the reporting of time of day information in the trials. Hence, in response, we have designed a standard report template to help integrate multiple oscillating physiological and pharmacokinetic components. Our hope is that by highlighting the need for accounting for temporal information when performing drug trials and measuring an aspect of physiology that displays diurnal variation, especially within this group of rare diseases, there is the potential for re-evaluating existing drugs which may not have been investigated at the most physiological relevant time of day.

## Data Availability

All data is included within the manuscript and supplemental table

## Acknowledgements

This work was supported by a generous grant from the Newnham College Senior Members Research Support fund.

## Author Contributions

L.S and T.H designed the study, L.S collated the data, L.S and T.H analysed the data, L.S and T.H wrote the paper, T.H conceived the idea

---

**Supplemental Table 1** – local database of clinical trials and inclusion criteria used in this study

